# Statistical Analysis Plan for the Aspiring to Reduce Events in Dialysis Trial (ASPIRED)

**DOI:** 10.64898/2026.01.12.26343774

**Authors:** Laurent Billot, Sarah Coggan, Li Fan, Vlado Perkovic, Xue Qing Yu, Sunil V. Badve, Meg Jardine, Tan Ning, Wei Chen, Helen Monaghan, Menghua Chen, Zhiming He, Guixian Ma, Sradha Kotwal, Muh Geot Wong

## Abstract

The ASPIRED trial aims to determine whether a daily low-dose of aspirin is effective in preventing major cardiovascular events when compared to placebo in patients receiving dialysis.

It is designed as a pragmatic trial two-arm double-blind randomised trial conducted within an established Dialysis Registry consisting of at least 80 centres and 30,000 participants.

This statistical analysis plan pre-specifies the method of analysis for every outcome and key variables collected in the trial. The primary outcome is time from randomisation to first occurrence of major cardiovascular event including myocardial infarction, ischemic stroke or death from cardiovascular cause.

The primary analysis will consist in a Cox proportional hazard model adjusted for stratification variables. The analysis plan also includes planned sensitivity analyses including covariate adjustments and subgroup analyses.

## 2 Administrative information

### 2.1 Study identifiers

- Protocol version: v6.0 05 Nov 2024
- ClinicalTrials.gov register Identifier: <u>NCT04381143</u>

### 2.2 Revision history

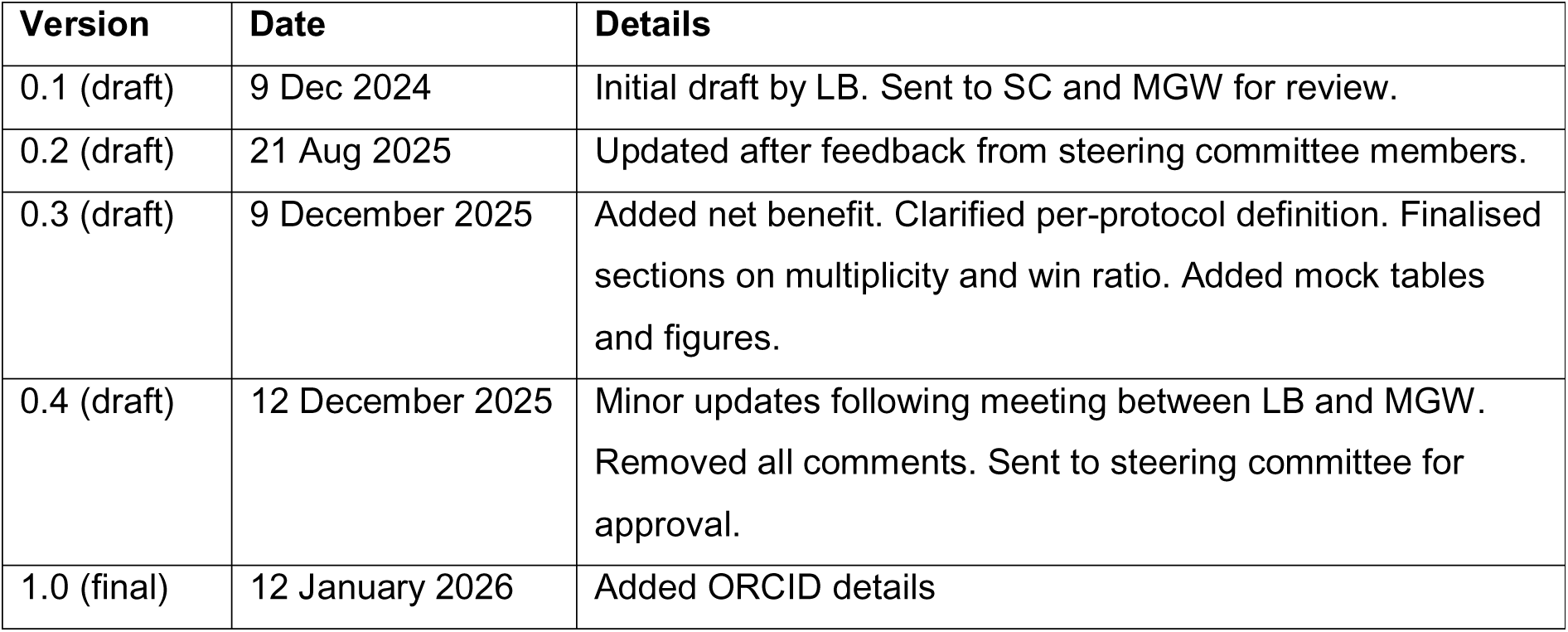
Causes of deaths

| Version | Date | Details |
| --- | --- | --- |
| 0.1 (draft) | 9 Dec 2024 | Initial draft by LB. Sent to SC and MGW for review. |
| 0.2 (draft) | 21 Aug 2025 | Updated after feedback from steering committee members. |
| 0.3 (draft) | 9 December 2025 | Added net benefit. Clarified per-protocol definition. Finalised sections on multiplicity and win ratio. Added mock tables and figures. |
| 0.4 (draft) | 12 December 2025 | Minor updates following meeting between LB and MGW. Removed all comments. Sent to steering committee for approval. |
| 1.0 (final) | 12 January 2026 | Added ORCID details |

### 2.3 Contributors to the statistical analysis plan

#### 2.2.1 Roles and responsibilities

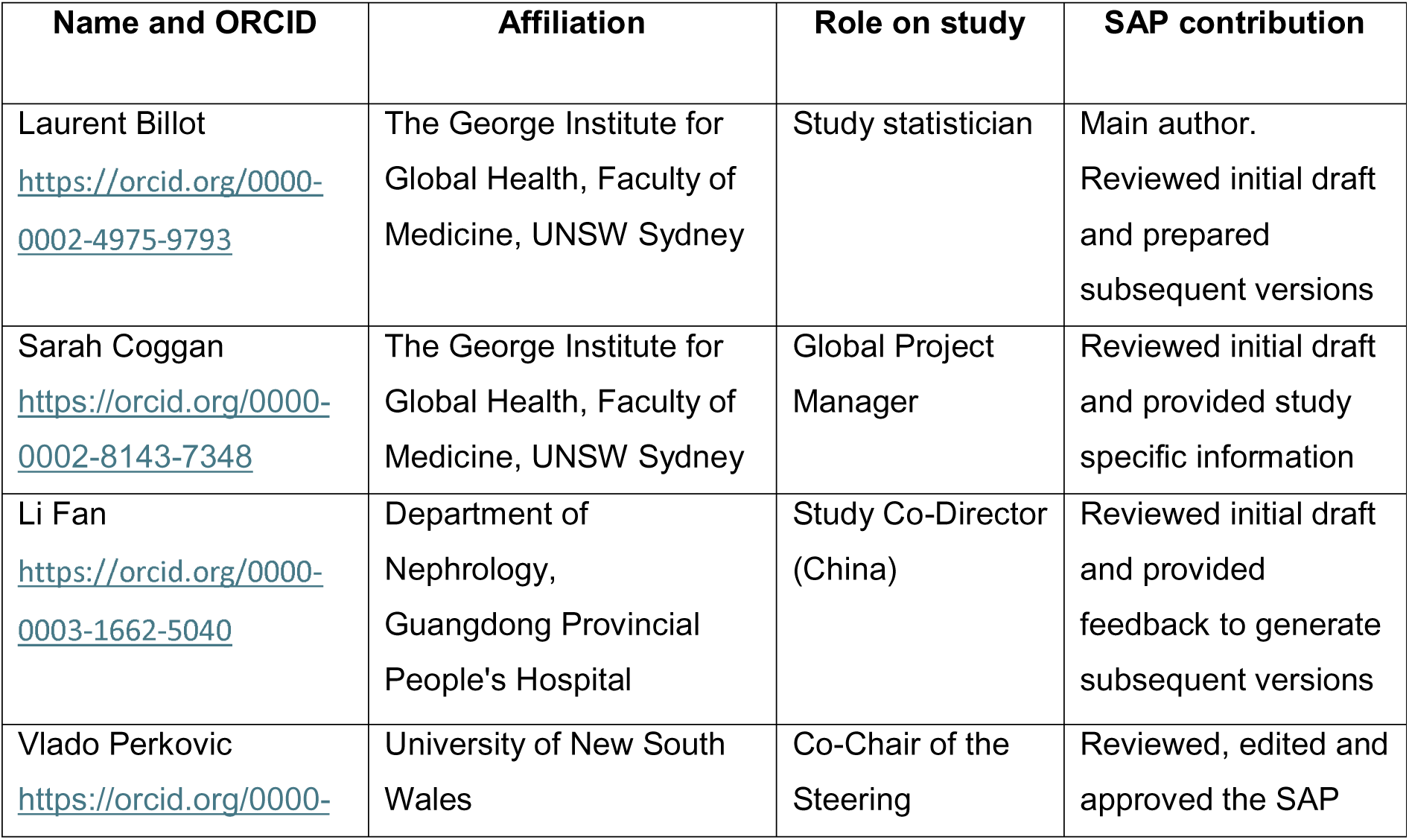
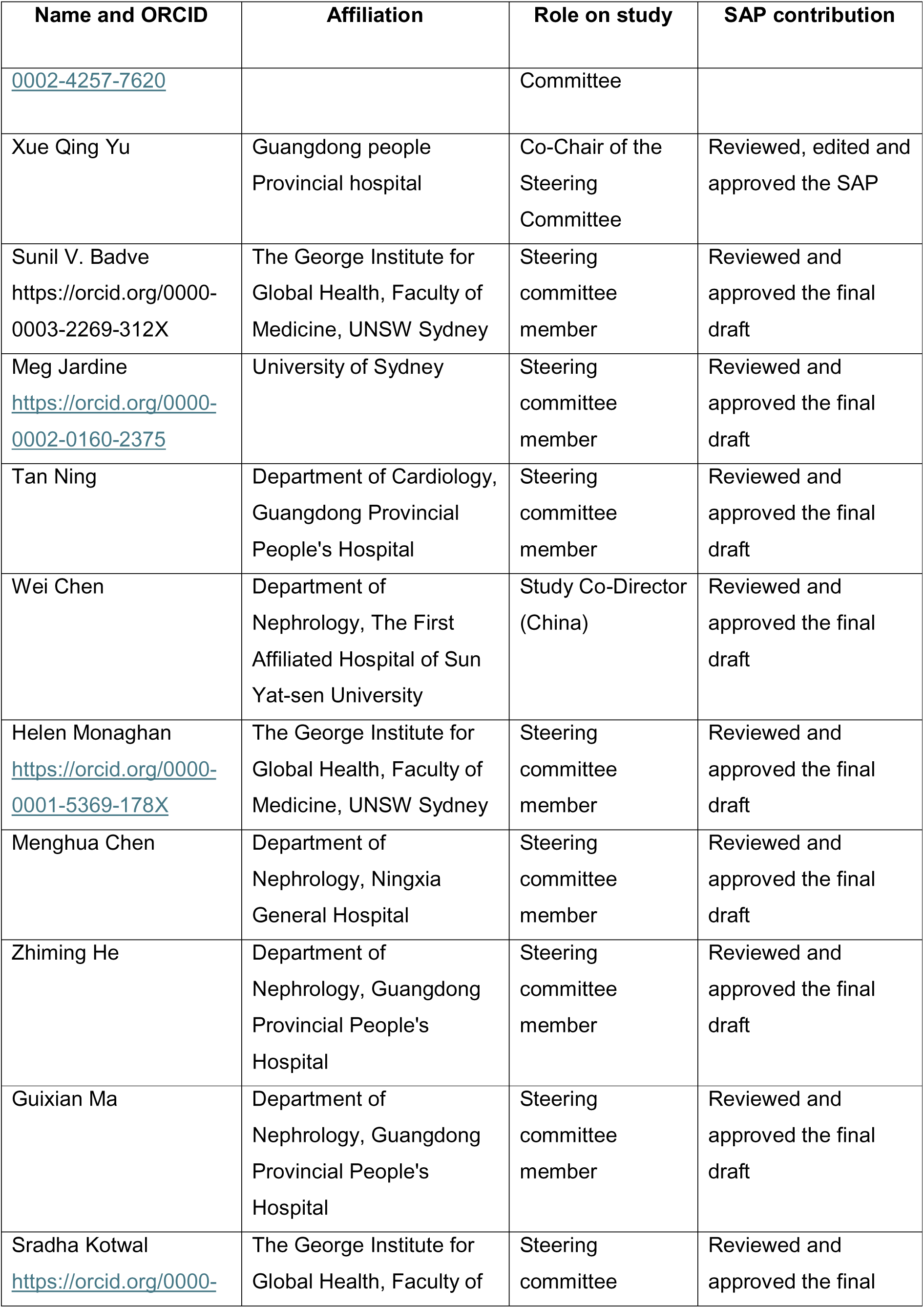
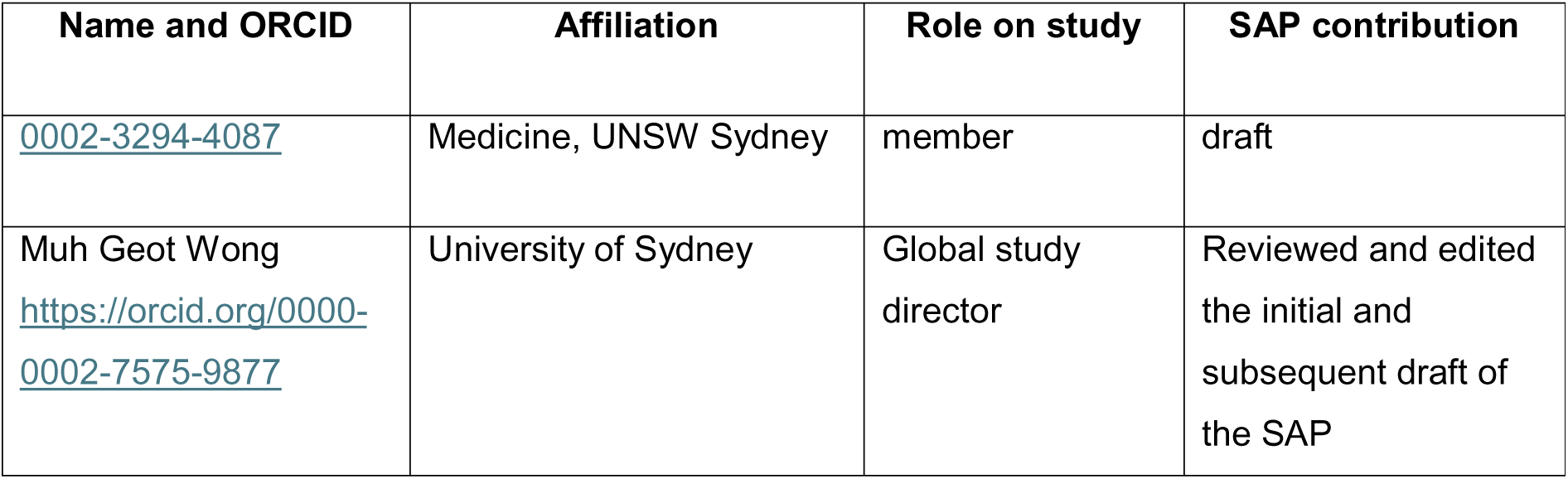
Causes of deaths

#### 2.2.2 Approvals

The undersigned have reviewed this plan and approve it as final. They find it to be consistent with the requirements of the protocol as it applies to their respective areas. They also find it to be compliant with International Conference on Harmonisation (ICH-E9) principles and in particular, confirm that this analysis plan was developed in a completely blinded manner (i.e. without knowledge of the effect of the intervention[s] being assessed).

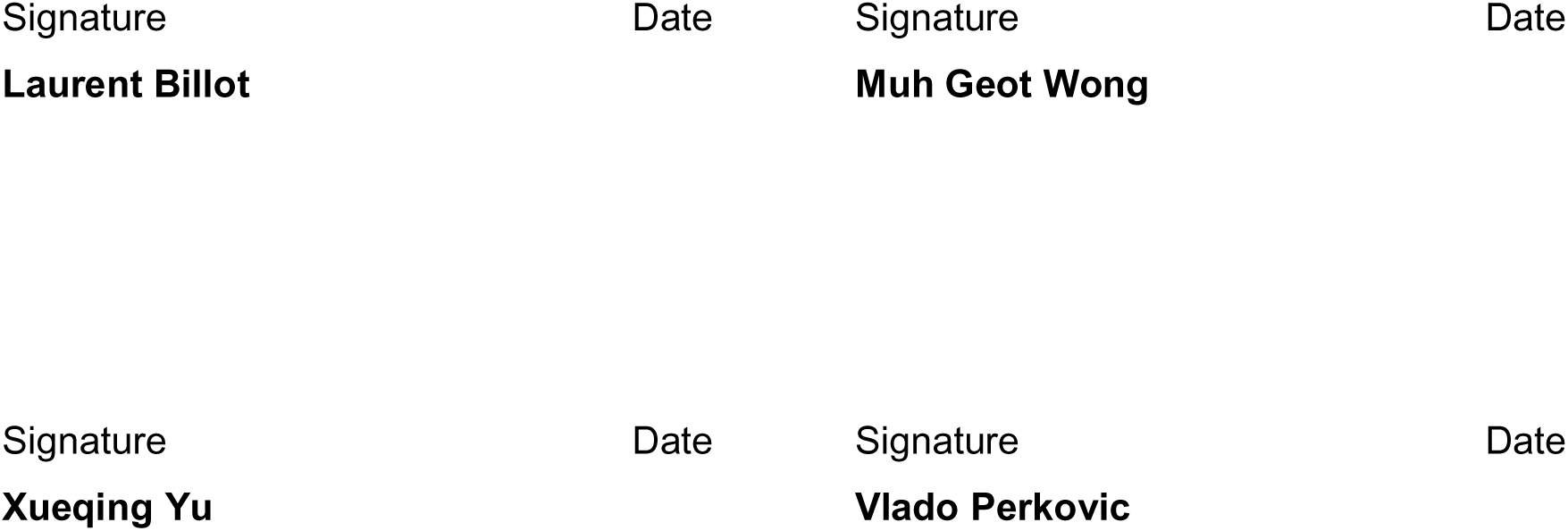
Causes of deaths

## 3 Introduction

### 3.1 Glossary of abbreviations and terms

Causes of deaths

| Abbreviation | Definition |
| --- | --- |
| AKI | Acute Kidney Injury |
| ASPIRED | ASPIrin in Reducing Events in Dialysis |
| BMI | Body Mass Index |
| BP | Blood pressure |
| CACE | complier average causal effect |
| CKD | Chronic Kidney Disease |
| CV | Cardiovascular |
| eGFR | Estimated Glomerular Filtration Rate |
| ESKD | End Stage Kidney Disease |
| HD | Haemodialysis |
| ISTH | International Society on Thrombosis and Haemostasis |
| ITT | Intention to Treat |
| MACE | Major Adverse Cardiac Events |
| MI | Myocardial Infarction |
| NOAC | new oral anticoagulant |
| PD | Peritoneal dialysis |
| PP | Per Protocol |
| SAE | Serious Adverse Event |
| SAP | Statistical Analysis Plan |
| TIA | Transient Ischemic Attack |
| UA | Unstable Angina |

### 3.2 Study synopsis

The ASPIrin in Reducing Events in Dialysis (ASPIRED) study is an investigator-initiated, pragmatic, double-blind, randomised controlled, clinical trial of ∼9,000 patients receiving dialysis (peritoneal dialysis (PD) or haemodialysis (HD)) to assess the efficiency and safety of low-dose aspirin (100 mg daily) versus placebo on prevention of major adverse cardiovascular events (myocardial infarction, ischemic stroke or death from cardiovascular cause).

### 3.3 Study population

This study is of pragmatic design conducted within an established Dialysis Registry in China, consisting of at least 80 centres and 30,000 existing patients receiving either PD or HD. Participants in this China Registry have provided informed consent to allow utilization of their Registry data. A separate HREC approved written consent will be obtained from eligible participants for the ASPIRED trial. A total of 9000 trial participants will be recruited from these sites with approximately thirty percent of the total trial participants are receiving peritoneal dialysis (PD) at the time of randomization.

#### 3.3.1 Eligibility/Inclusion Criteria

1. Incident or prevalent adults (≥18 years old) dialysis patients are included in the Dialysis Registry
2. Commenced on dialysis with the expectation of ongoing maintenance dialysis requirement
3. Willing and able to provide informed consent for this study

#### 3.3.2 Exclusion criteria

1. Requirement for any form of antiplatelet agent (aspirin, glycoprotein IIb/IIIa inhibitors etc), or oral anticoagulation (warfarin, new oral anticoagulants (NOACs)), in the view of the treating physician
2. Contraindication to aspirin, in the view of the treating physician
3. Dialysis requirement due to acute kidney injury (AKI) with expectation of kidney function recovery
4. History of haemorrhagic stroke or intracranial bleed within 12 months of screening
5. Coagulopathy from any cause
6. Unable to provide informed consent

### 3.4 Interventions

Consented participants are randomly allocated to either oral aspirin 100 mg daily, or placebo. Participants will be advised to take this medication per manufacture’s instruction. The investigator is permitted to interrupt study drug for a given participant if continuation is deemed to be detrimental to the participant’s well-being. All participants who interrupt study drug should resume treatment when possible, as long as it is considered safe to do so. If there is concern that the participant may be intolerant of study treatments or if the participant is reluctant to take the full dose of study treatments, a pragmatic approach is adopted allowing restarting study medications at a reduced frequency e.g. alternate-daily, as deemed appropriate by the investigators Irrespective of whether or not treatment is resumed, all participants are encouraged to continue protocol defined study follow up until the end of the study.

### 3.5 Outcomes

#### 3.5.1 Primary endpoint

The primary endpoint is a major adverse cardiovascular events composite (MACE), defined as the time from randomisation first occurrence of myocardial infarction (MI), ischemic stroke or death from cardiovascular causes.

#### 3.5.2 Primary safety endpoint

The primary safety variable will be the time from randomisation to the first occurrence of major bleeding based on the modified International Society on Thrombosis and Haemostasis (ISTH) major bleeding definition i.e.:

- fatal bleeding, and/or
- symptomatic bleeding in a critical area or organ, such as intracranial, intra-spinal, intra-ocular, retro-peritoneal, intra-articular or pericardial, or intramuscular with compartment syndrome, or bleeding into a surgical site requiring re-operation and/or
- bleeding leading to hospitalization

#### 3.5.3 Secondary endpoints

Secondary endpoints include the following:

1. Composite of major cardiovascular events (defined above) and all-cause death
2. Composite of major cardiovascular events, hospitalised unstable angina (UA) and hospitalised transient ischaemic attack (TIA)
3. Individual components of the composite
4. All-cause mortality
5. Coronary revascularisation
6. Fistula or graft thrombosis
7. Intracranial haemorrhage
8. Peripheral vascular events
9. Ischemic events (defined as a composite of MI, death due to cardiovascular cause, ischemic stroke, or any revascularization procedure (i.e. exclusion of deaths and strokes confirmed to be haemorrhagic))

### 3.6 Randomisation and blinding

Consented patients are randomised using a web-based system via a password- protected encrypted website to either low dose aspirin 100mg daily, or placebo, in a 1:1 ratio. A covariate-balancing adaptive allocation algorithm is used to minimise imbalance across treatment groups in the following variables: (1) site, (2) modality of dialysis (PD or HD), (3) previous history of cardiovascular disease, and (4) presence or absence of diabetes mellitus. Trial participants, site staff and study team members will be blinded to an individual’s randomised allocation

### 3.7 Statistical hypotheses

We hypothesise that the use of low dose aspirin will reduce the incidence of major cardiovascular events safely in people with stage V CKD on maintenance dialysis.

### 3.8 Sample size

Assuming a primary endpoint event rate of 4 per 100 person-years in the control group and a two-sided log-rank test with an alpha of 5%, a sample size of 9,000 participants will provide 90% power to detect a 18.5% relative risk reduction (HR=0.815), or 80% power to detect a 16.2% relative risk reduction (HR=0.838) with low dose aspirin versus placebo. This corresponds to approximately 1011 participants experiencing a primary endpoint event.

We anticipate 60 months of recruitment will be needed to enrol 9,000 patients. A fixed design without interim analysis would require 17 months of follow-up after the last patient is recruited (total duration of 77 months). We are planning 2-3 interim efficacy analysis which are expected to reduce follow-up to 8-9 months under the alternative hypothesis (total duration of 68-69 months).

## 4 Statistical analyses

### 4.1 General principles

#### 4.1.1 Level of statistical significance

Up to a total of 3 interim analyses are planned using Haybittle-Peto boundaries for efficacy [1]. The final significance level for the analysis of the primary outcome will be adjusted based on the exact number and timing of interim analyses conducted to maintain the overall type-I error rate at 5% (two-sided). These will be calculated using RPACT [2] based on the number of events available at the time of each interim analysis to determine the information fraction.

Given the clear outcome hierarchy, the analysis of the primary outcome will not be adjusted for multiplicity (aside from the one potentially arising from interim analyses). Sensitivity analyses of the primary outcome (e.g. additional covariate adjustments or per-protocol analyses) will also not adjust for multiplicity as they are conducted to assess the robustness of the main findings using different approaches/assumptions.

For the secondary outcomes, we will use the following approach:

1. For all-cause death and the three components of the primary outcome (cardiovascular death, ischemic stroke and myocardial infarction), the family-wise error rate will be controlled using a hierarchical step-down approach with a Holm-Sidak correction [3]. Briefly, the Holm-Sidak approach consists of ordering all p-values from smallest to largest, and then comparing them to an adjusted level of significance calculated as 1-(1-0.05)^1/C^, where C indicates the number of comparisons that remain. With 4 endpoints, the smallest p value will be compared to 1-(1-0.05)^1/4^, the second p value to 1-(1-0.05)^1/3^, with the last one being compared to 1-(1-0.05) (i.e. 0.05). The sequential testing procedure stops as soon as a p value fails to reach the corrected significance level.
2. For the remaining secondary outcomes including the alternative composite outcomes, no formal test will be conducted and we will only report point estimates and 95% confidence intervals to assess consistency of treatment effects.

For other outcomes including safety outcomes and other analyses (e.g. win ratio or net clinical benefit) we will not adjust the significance level which will be kept at 5% given it includes key safety outcomes and we wish to be able to detect a potential signal.

#### 4.1.2 Analysis populations

All analyses will be performed in the intention to treat (ITT) analysis set. Sensitivity analyses for selected outcomes (as flagged in the relevant sections of the SAP) will be conducted after adjusting for compliance including per-protocol and/or complier average causal effect (CACE) analyses.

The ***intention-to-treat (ITT) analysis set*** will include all randomised eligible participants. Participants will be analysed according to the group they were randomised to, regardless of treatment adherence or post-randomisation events. The ITT set will be used to assess all efficacy and safety outcomes.

The ***per-protocol (PP) analysis set*** will be defined as patients who are still taking the study drug at the time of the last visit i.e. answered “yes” to the Question “Is the participant still taking randomized treatment?”) regardless of potential temporary discontinuations. The PP set will be used for sensitivity analyses of the primary outcome.

Post-hoc **CACE analyses** may be conducted depending on the main results. They are not described in detail in this SAP.

#### 4.1.3 Missing data handling

Except where specifically indicated, analyses will be conducted using all available data with no imputation. Given the nature of the primary endpoint (time to major cardiovascular outcome), there is no reasonable way to assess whether some events are potentially missing (i.e. unreported). We will therefore perform the analysis assuming that all events that occurred were reported, and without imputation of missing data. All reported events will be included in the time-to-first-event analysis with every participant censored at the known time of event, or the time when the participant was last known to be alive and free of an event. The same approach will be used for every survival endpoint.

#### 4.1.4 Adjustment for stratification variables

Randomisation is stratified by four variables: study site, modality of dialysis (PD or HD), previous history of cardiovascular disease and presence or absence of diabetes mellitus. By default, all analyses will be adjusted by these four variables with site as a random effect and the three others as fixed effects. Further covariate adjustments will be specifically indicated where relevant.

#### 4.1.5 Statistical software

Analyses will be conducted primarily using SAS (version 9.3 or above) or R (version R-4.3.1 or above).

### 4.2 Analysis of baseline data

Socio-demographic characteristics, medical history, concomitant medications and laboratory data at the time of randomisation (baseline) will be summarised by treatment arm and overall using standard summary statistics i.e. n, mean, standard-deviation, median, quartiles, minimum and maximum for continuous variables and numerator, denominator and percentage for categorical variables. No statistical test will be performed.

### 4.3 Analysis of laboratory data and vital signs

Laboratory data and vital signs will be summarised descriptively by treatment arm and visit using tables and longitudinal plots showing means and 95% confidence intervals. No formal modelling or statistical test will be performed.

### 4.4 Analysis of compliance

Medication compliance will be reported using the following metrics:

- Proportion of patients still on randomised therapy by visit
- Number and percentage of discontinuations by type (permanent or temporary) and reason
- Protocol deviations

These will be analysed descriptively only.

### 4.5 Analysis of the primary outcome

The primary outcome is a composite of myocardial infarction, ischemic stroke or death from cardiovascular causes. The main analysis will consist of a survival analysis of time to first event conducted with a Cox proportional model. Participants will be censored at the time when they receive kidney transplantation or die from a cause other than cardiovascular. Censoring will otherwise occur when the participant was last known to be alive and free of event. As a sensitivity analysis, kidney transplantation and death from non-cardiovascular causes will be treated as competing risks (see Section 4.5.2.2 for details).

#### 4.5.1 Main analysis of the primary outcome

Survival will be described using Kaplan-Meier curves of time-to-first event. The effect of the intervention will be estimated as a hazard ratio and confidence interval from a Cox model. To model potential within-site correlations due to stratification, we will use a shared-parameter frailty Cox model with a random site effect [4]. Fixed covariates will include the randomised treatment and the other 3 stratification variables (dialysis modality, cardiovascular disease history and presence of diabetes mellitus).

#### 4.5.2 Sensitivity analyses

##### 4.5.2.1 Adjusted analyses

The Cox model described in Section 4.5.1 will be rerun after adding the following baseline covariates to the model: age (as a continuous variable), sex, smoking status, diabetes and dialysis vintage (less than 3 years, 3-5 years, more than 5 years).

##### 4.5.2.1 Competing risk analysis

The model described in Section 4.5.1 will be rerun by treating kidney transplantation and death from non-cardiovascular causes as competing risks. Survival will be summarized using cumulative incidence functions and the effect of the intervention will be derived from a Fine-Gray model as the sub-distribution hazard ratio and confidence interval [5].

##### 4.5.2.1 Subgroup analyses

The following subgroup analyses are planned for the primary outcome, regardless of the statistical significance for the primary analysis:

- Age: <40, 40-<50, 50-<60, 60+
- Sex: male vs. female
- Dialysis modality: HD vs PD
- History of cardiovascular disease: yes vs no
- Diabetes mellitus: yes vs no
- Smoking status: smoker vs non-smoker

### 4.6 Analysis of secondary and safety outcomes

#### 4.6.1 Survival analyses

Each secondary outcome will be analysed using the same approach as the primary outcome described in Section 4.5.1 i.e. described using KM plots and using a Cox model of time to first event with adjustment for stratification variables to estimate the effect of the intervention. Except where the outcome itself is or includes all-cause mortality, death will be treated as a censoring mechanism. Participants will also be censored at the time of kidney transplantation.

#### 4.6.2 Safety analyses

Major bleeding events will be summarised as the number of events as well as the number and proportion of patients experiencing at least one event. This will be done overall, by outcome and by site of bleeding.

Time to first major bleeding event will be described by randomised arm using Kaplan-Meier plots and analysed using a Cox model adjusted for stratification variables using the same approach as the analysis of the primary efficacy outcome (see Section 4.5.1). Censoring will occur when the patient was last known to be alive and free of major bleeding event.

We will separately report the number and proportion of patients with colorectal cancer as an adverse event of special interest.

These analyses will be repeated in the per-protocol analysis population.

#### 4.6.3 Net Benefit analysis

To assess the overall (net) benefit balance of the intervention, we will analyse a composite of major efficacy and safety outcomes defined as a composite of cardiovascular death, ischemic stroke, myocardial infarction, hospitalisation for unstable angina or transient ischaemic attack, coronary revascularisation, peripheral vascular event, fistula or graft thrombosis and major bleeding events. Time to first event will be described by randomised arm using Kaplan-Meier plots and analysed using a Cox model adjusted for stratification variables using the same approach as the analysis of the primary efficacy outcome (see Section 4.5.1). Censoring will occur when the patient was last known to be alive and free of event. These analyses will be repeated in the per-protocol analysis population

#### 4.6.4 Win ratio analysis

All components of the primary, secondary and key safety outcomes will be analysed jointly using a win ratio approach which ranks all outcomes in order of importance [6]. The following hierarchy will be used:

1. Cardiovascular death
2. Ischemic stroke
3. Myocardial infarction
4. Hospitalised for unstable angina or transient ischaemic attack
5. Coronary revascularisation
6. Peripheral vascular events
7. Fistula or graft thrombosis

Every participant randomised to the aspirin arm will be paired with every participant randomised to the placebo arm. For illustrative purposes, if we had 130 patients randomised to one arm and 127 to the other, we would compare a total of 16,510 pairs (130 × 127). Within each pair, we will compare outcomes in a hierarchical fashion until a winner has been determined. If no winner can be declared after comparing all outcomes, the pair will be tied. We will start by comparing cardiovascular mortality to determine a “winner”. The winner will be the participant with the longest survival time. If neither participant died and a winner cannot be declared on mortality alone, we will then proceed to the next step and compare time to stroke. As with death, the winner will be the participant with the longest time to first event. If neither patient experienced the event of interest or both experienced the event on the same day, the pair will be declared a tie and we will proceed to the next outcome in the hierarchy. Only events that occur during the time that both participants are in the study will be used (e.g. if one participant withdraws after 1 year, only the events occurring in the first year will be used to compare the pair). The approach will proceed in a stepwise fashion, moving to the next outcome in the hierarchy, until a decision has been reached for every pair. The full decision process is described in the table below. The win ratio will then be calculated as the proportion of “aspirin winners” (the participant in the aspirin arm had a better outcome) divided by the proportion of “placebo winners” (the participant in the placebo arm had a better outcome)

To complete the interpretation, we will also compute the win odds and net benefit [7]. Calculations will be performed using the R package WINS developed by Cui and Huang [8].

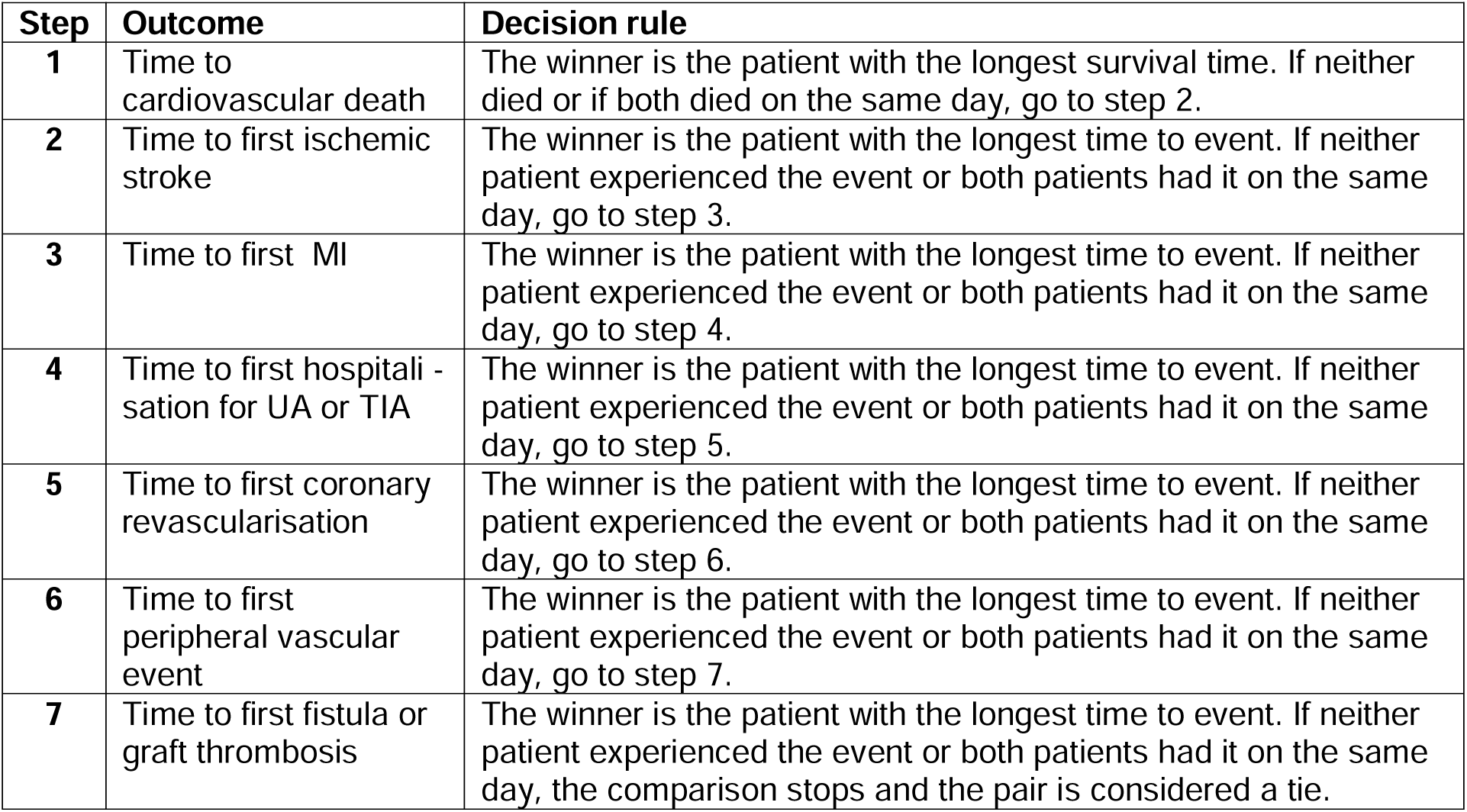
Causes of deaths

An “incremental analysis” will also be performed by calculating the win ratio and 95% CI after adding each outcome i.e. for the first outcome only, for the first two together, the first three, and so on (cf Figure 5 for details).

**Figure 1.**
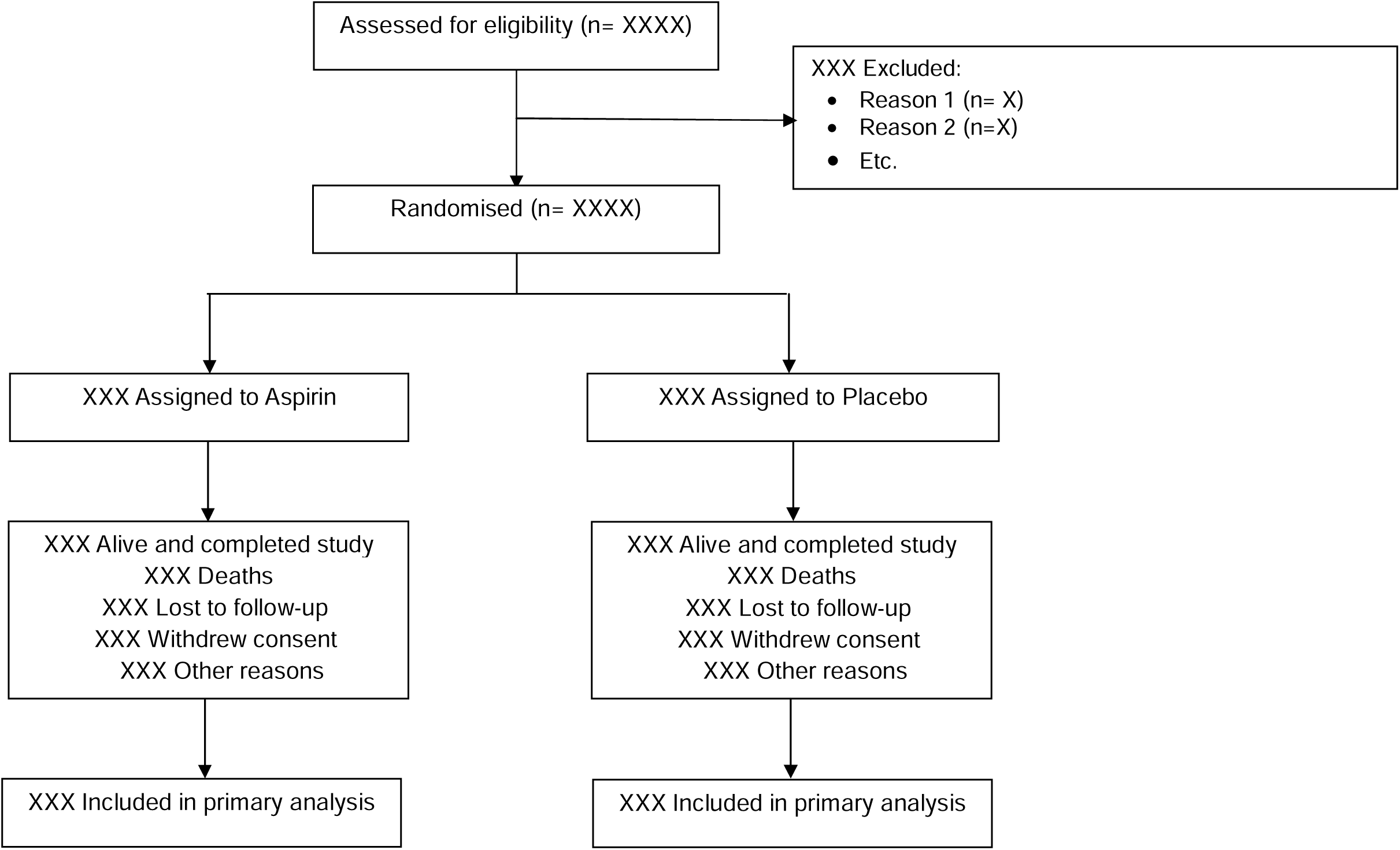
**Consort flow chart**

**Figure 2.**
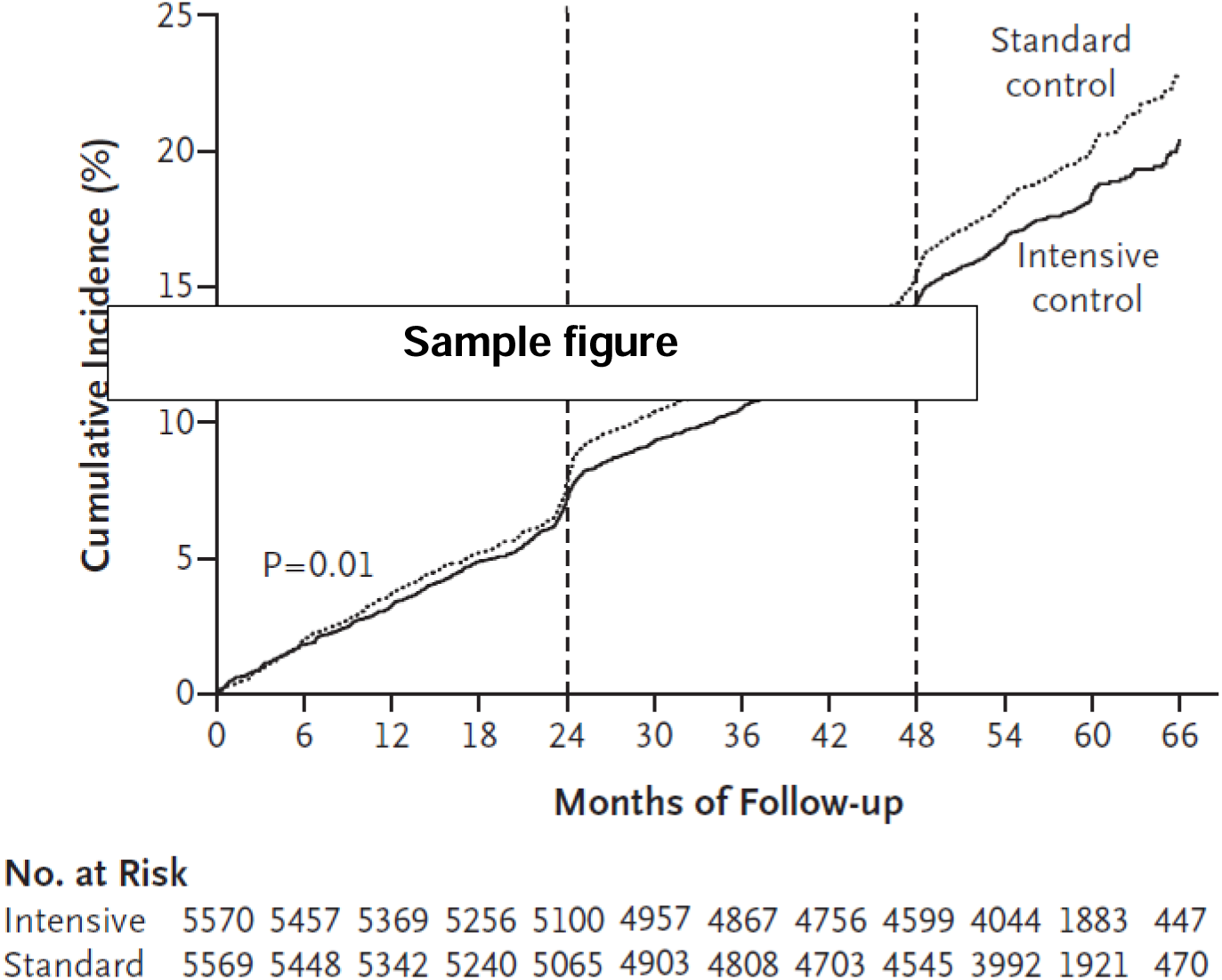
Kaplan-Meier plot of time to first primary outcome Figure above for illustrative purposes only (source: https://www.nejm.org/doi/full/10.1056/NEJMoa0802987) Notes: do for every survival outcome. Show number at risk every 6 months. Display hazard ratio, 95% CI and p-value obtained from the Cox model.

**Figure 3.**
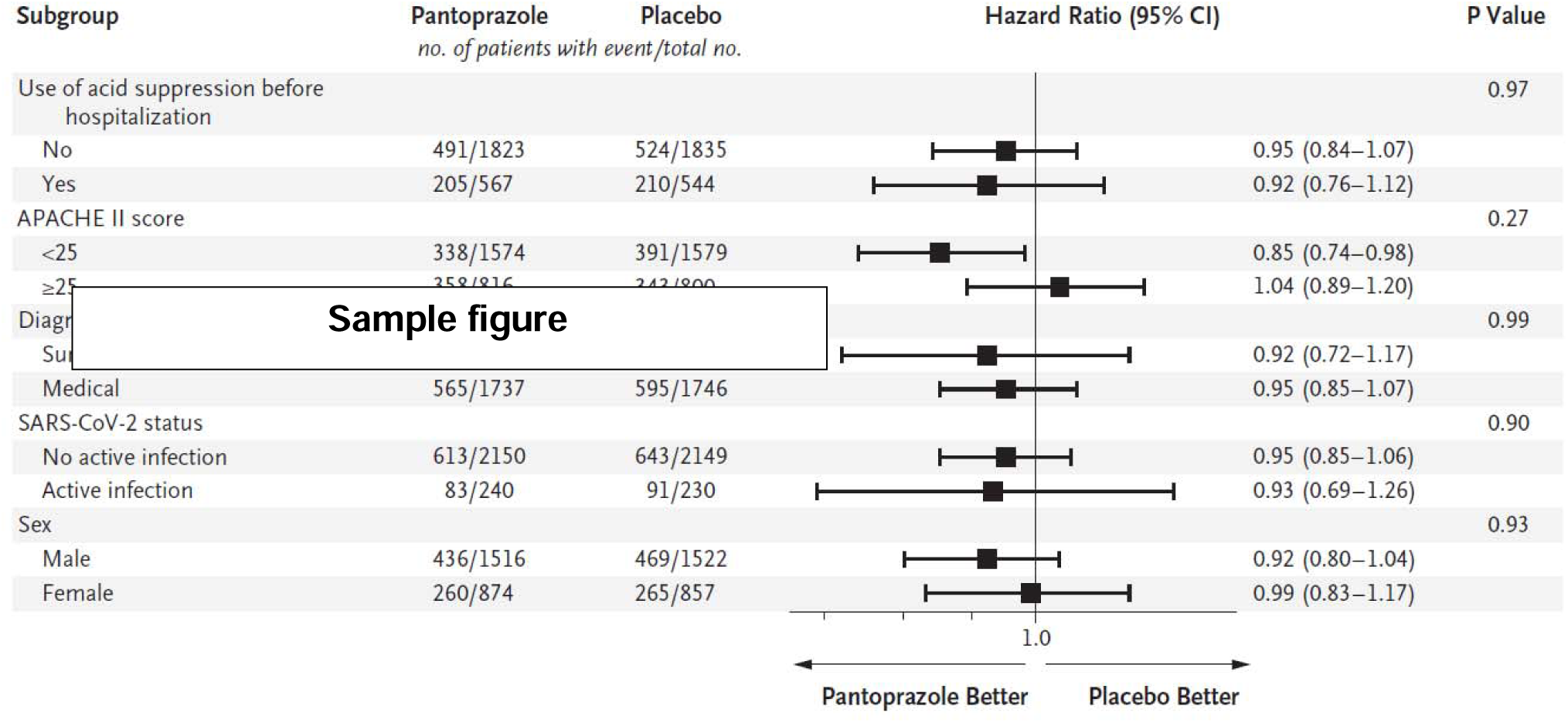
Forest plot of subgroup analyses Notes: Do for the primary outcome only. Figure above for illustrative purposes only (source: https://www.nejm.org/doi/abs/10.1056/NEJMoa2404245)

**Figure 4.**
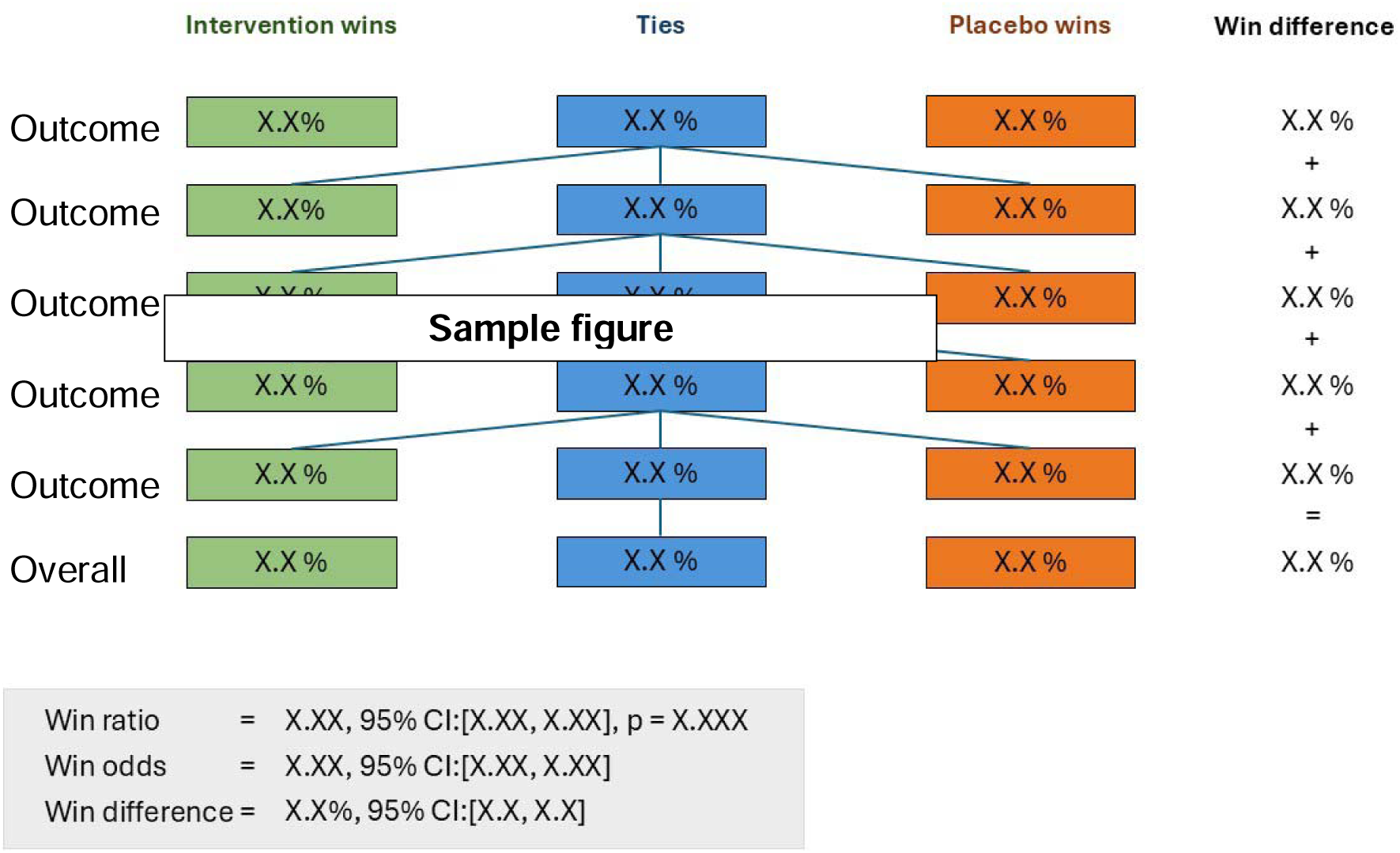
Win ratio decision tree

**Figure 5.**
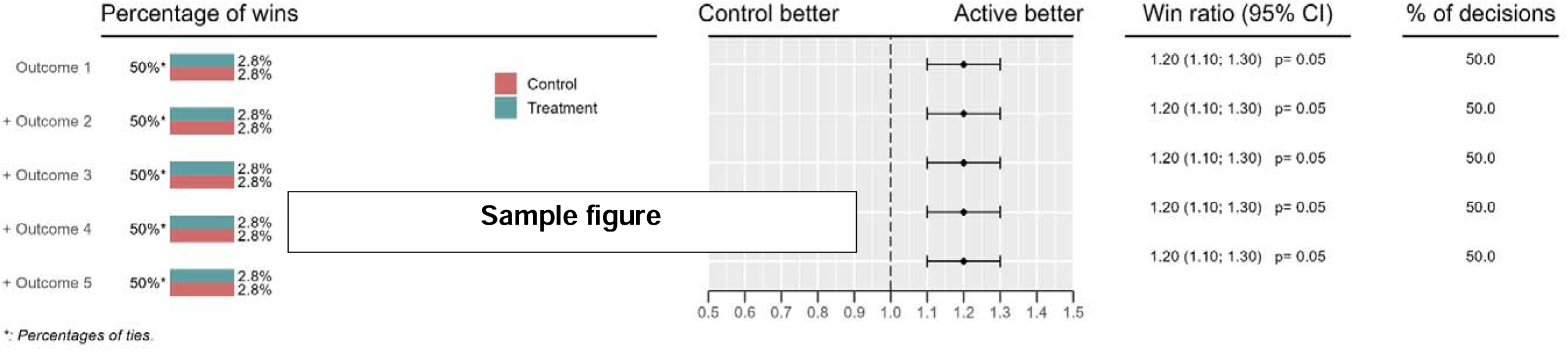
Win ratio cumulative analysis

**Figure 6.** Lab results and vitals over time Note: longitudinal plots showing means and 95% confidence intervals by treatment and by visit. Display the number of patients with available data at every visit. Create this plot for every lab parameter and vital sign collected over time.

## 6 Planned key outputs

**Table 1.** Participant disposition.

|  | **Aspirin**<br>**(N = XXX)** | **Placebo**<br>**(N = XXX)** | **Total**<br>**(N = XXX)** |
| --- | --- | --- | --- |
| **Visit attendance** |  |  |  |
| Month 3 | XX (XX%) | XX (XX%) | XX (XX%) |
| Month 6 | XX (XX%) | XX (XX%) | XX (XX%) |
| Month 12 | XX (XX%) | XX (XX%) | XX (XX%) |
| Month 18 | XX (XX%) | XX (XX%) | XX (XX%) |
| Etc. |  |  |  |
| **Trial withdrawal** |  |  |  |
| Reason 1 | XX (XX%) | XX (XX%) | XX (XX%) |
| Reason 2 | XX (XX%) | XX (XX%) | XX (XX%) |
| Etc. | XX (XX%) | XX (XX%) | XX (XX%) |

*Note: the denominator is all randomised patients*

**Table 2.**
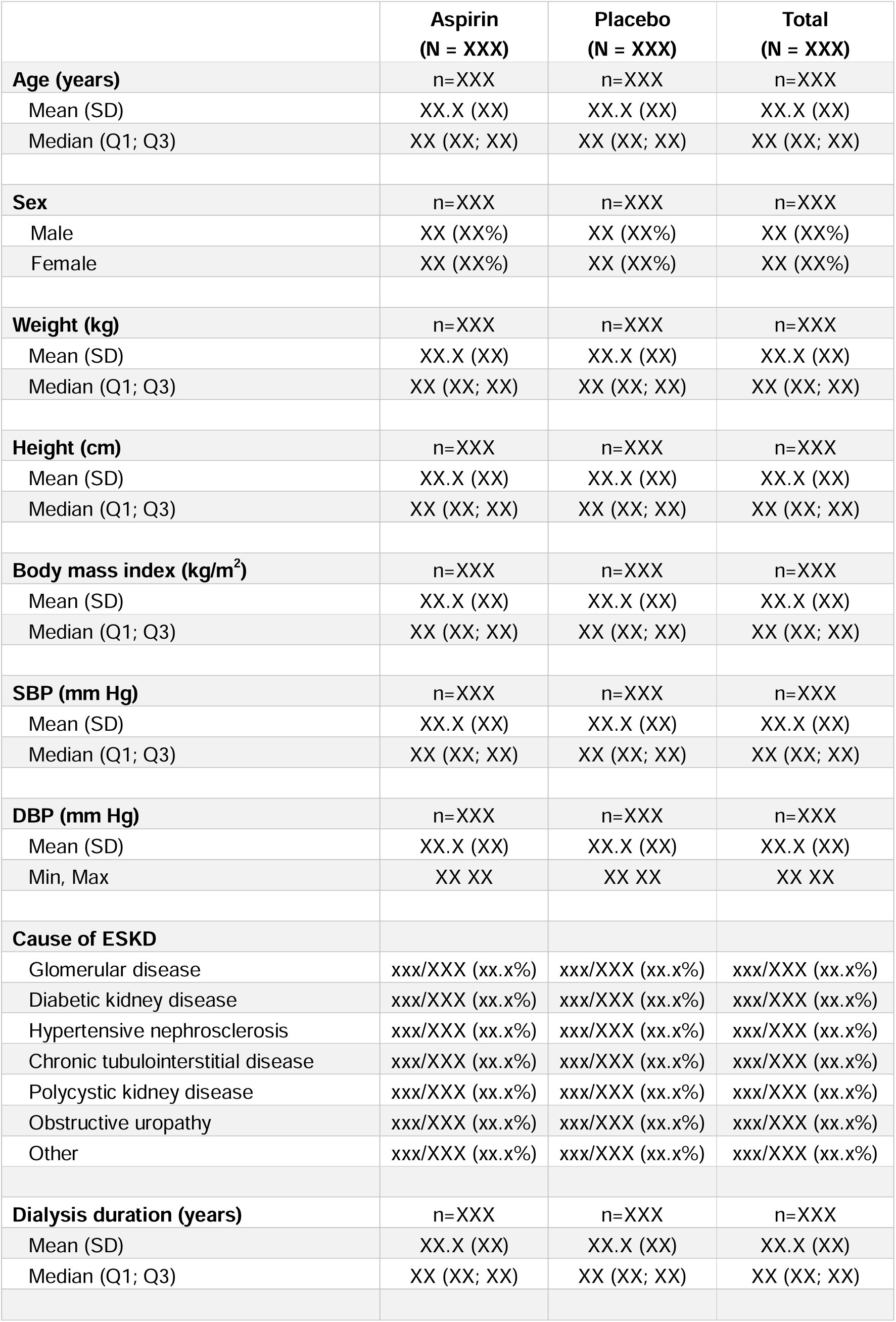
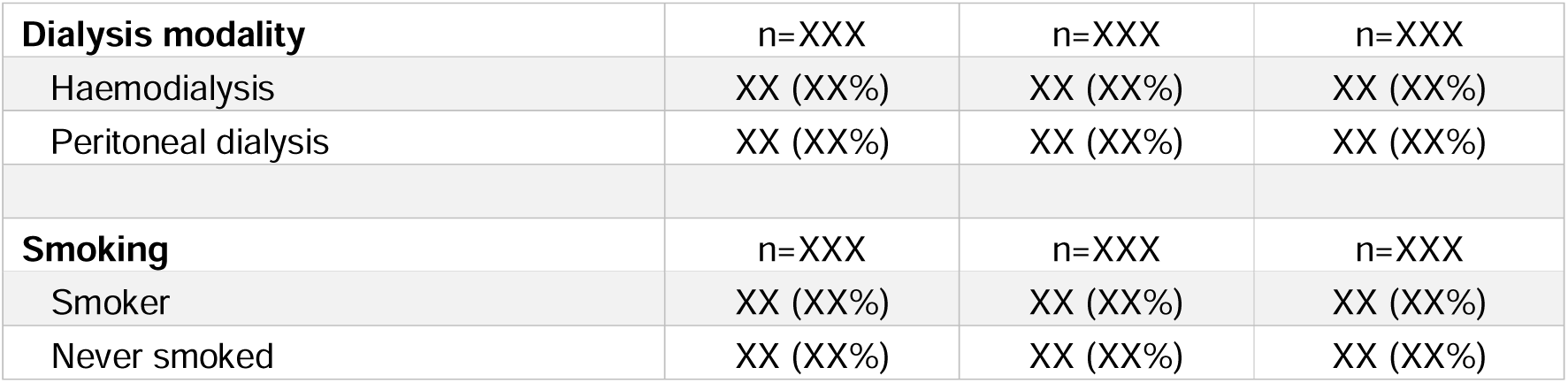
Key baseline characteristics.

|  | **Aspirin**<br>**(N = XXX)** | **Placebo**<br>**(N = XXX)** | **Total**<br>**(N = XXX)** |
| --- | --- | --- | --- |
| **Age (years)** | n=XXX | n=XXX | n=XXX |
| Mean (SD) | XX.X (XX) | XX.X (XX) | XX.X (XX) |
| Median (Q1; Q3) | XX (XX; XX) | XX (XX; XX) | XX (XX; XX) |
| **Sex** | n=XXX | n=XXX | n=XXX |
| Male | XX (XX%) | XX (XX%) | XX (XX%) |
| Female | XX (XX%) | XX (XX%) | XX (XX%) |
| **Weight (kg)** | n=XXX | n=XXX | n=XXX |
| Mean (SD) | XX.X (XX) | XX.X (XX) | XX.X (XX) |
| Median (Q1; Q3) | XX (XX; XX) | XX (XX; XX) | XX (XX; XX) |
| **Height (cm)** | n=XXX | n=XXX | n=XXX |
| Mean (SD) | XX.X (XX) | XX.X (XX) | XX.X (XX) |
| Median (Q1; Q3) | XX (XX; XX) | XX (XX; XX) | XX (XX; XX) |
| **Body mass index (kg/m<sup>2</sup>)** | n=XXX | n=XXX | n=XXX |
| Mean (SD) | XX.X (XX) | XX.X (XX) | XX.X (XX) |
| Median (Q1; Q3) | XX (XX; XX) | XX (XX; XX) | XX (XX; XX) |
| **SBP (mm Hg)** | n=XXX | n=XXX | n=XXX |
| Mean (SD) | XX.X (XX) | XX.X (XX) | XX.X (XX) |
| Median (Q1; Q3) | XX (XX; XX) | XX (XX; XX) | XX (XX; XX) |
| **DBP (mm Hg)** | n=XXX | n=XXX | n=XXX |
| Mean (SD) | XX.X (XX) | XX.X (XX) | XX.X (XX) |
| Min, Max | XX XX | XX XX | XX XX |
| **Cause of ESKD** |  |  |  |
| Glomerular disease | xxx/XXX (xx.x%) | xxx/XXX (xx.x%) | xxx/XXX (xx.x%) |
| Diabetic kidney disease | xxx/XXX (xx.x%) | xxx/XXX (xx.x%) | xxx/XXX (xx.x%) |
| Hypertensive nephrosclerosis | xxx/XXX (xx.x%) | xxx/XXX (xx.x%) | xxx/XXX (xx.x%) |
| Chronic tubulointerstitial disease | xxx/XXX (xx.x%) | xxx/XXX (xx.x%) | xxx/XXX (xx.x%) |
| Polycystic kidney disease | xxx/XXX (xx.x%) | xxx/XXX (xx.x%) | xxx/XXX (xx.x%) |
| Obstructive uropathy | xxx/XXX (xx.x%) | xxx/XXX (xx.x%) | xxx/XXX (xx.x%) |
| Other | xxx/XXX (xx.x%) | xxx/XXX (xx.x%) | xxx/XXX (xx.x%) |
| **Dialysis duration (years)** | n=XXX | n=XXX | n=XXX |
| Mean (SD) | XX.X (XX) | XX.X (XX) | XX.X (XX) |
| Median (Q1; Q3) | XX (XX; XX) | XX (XX; XX) | XX (XX; XX) |
| **Dialysis modality** | n=XXX | n=XXX | n=XXX |
| Haemodialysis | XX (XX%) | XX (XX%) | XX (XX%) |
| Peritoneal dialysis | XX (XX%) | XX (XX%) | XX (XX%) |
| **Smoking** | n=XXX | n=XXX | n=XXX |
| Smoker | XX (XX%) | XX (XX%) | XX (XX%) |
| Never smoked | XX (XX%) | XX (XX%) | XX (XX%) |

**Table 3.** Medical history and concomitant medications at baseline.

|  | **Aspirin**<br>**(N = XX)** | **Placebo**<br>**(N = XXX)** | **All**<br>**(N = XX)** |
| --- | --- | --- | --- |
| **Comorbidities** |  |  |  |
| Cardiovascular disease (defined as patient had coronary artery disease or cerebral infarction) | xxx/XXX (xx.x%) | xxx/XXX (xx.x%) | xxx/XXX (xx.x%) |
| Intracranial haemorrhage | xxx/XXX (xx.x%) | xxx/XXX (xx.x%) | xxx/XXX (xx.x%) |
| Diabetes | xxx/XXX (xx.x%) | xxx/XXX (xx.x%) | xxx/XXX (xx.x%) |
| Heart failure | xxx/XXX (xx.x%) | xxx/XXX (xx.x%) | xxx/XXX (xx.x%) |
| Arrhythmia | xxx/XXX (xx.x%) | xxx/XXX (xx.x%) | xxx/XXX (xx.x%) |
| Peripheral vascular disease | xxx/XXX (xx.x%) | xxx/XXX (xx.x%) | xxx/XXX (xx.x%) |
| Hypertension | xxx/XXX (xx.x%) | xxx/XXX (xx.x%) | xxx/XXX (xx.x%) |
| Hyperlipidemia | xxx/XXX (xx.x%) | xxx/XXX (xx.x%) | xxx/XXX (xx.x%) |
| Peptic ulcer/gastrointestinal bleeding | xxx/XXX (xx.x%) | xxx/XXX (xx.x%) | xxx/XXX (xx.x%) |
| Colorectal cancer | xxx/XXX (xx.x%) | xxx/XXX (xx.x%) | xxx/XXX (xx.x%) |
| History of kidney transplantation | xxx/XXX (xx.x%) | xxx/XXX (xx.x%) | xxx/XXX (xx.x%) |
| **Medications** |  |  |  |
| Erythropoietin | xxx/XXX (xx.x%) | xxx/XXX (xx.x%) | xxx/XXX (xx.x%) |
| HIF-PHI inhibitors | xxx/XXX (xx.x%) | xxx/XXX (xx.x%) | xxx/XXX (xx.x%) |
| Iron agent | xxx/XXX (xx.x%) | xxx/XXX (xx.x%) | xxx/XXX (xx.x%) |
| Phosphorus binder | xxx/XXX (xx.x%) | xxx/XXX (xx.x%) | xxx/XXX (xx.x%) |
| Lipid-lowering agents | xxx/XXX (xx.x%) | xxx/XXX (xx.x%) | xxx/XXX (xx.x%) |
| RAS inhibitors | xxx/XXX (xx.x%) | xxx/XXX (xx.x%) | xxx/XXX (xx.x%) |
| Beta-blocker | xxx/XXX (xx.x%) | xxx/XXX (xx.x%) | xxx/XXX (xx.x%) |
| Proton-pump inhibitor | xxx/XXX (xx.x%) | xxx/XXX (xx.x%) | xxx/XXX (xx.x%) |
| H2 antagonists | xxx/XXX (xx.x%) | xxx/XXX (xx.x%) | xxx/XXX (xx.x%) |

**Table 4.**
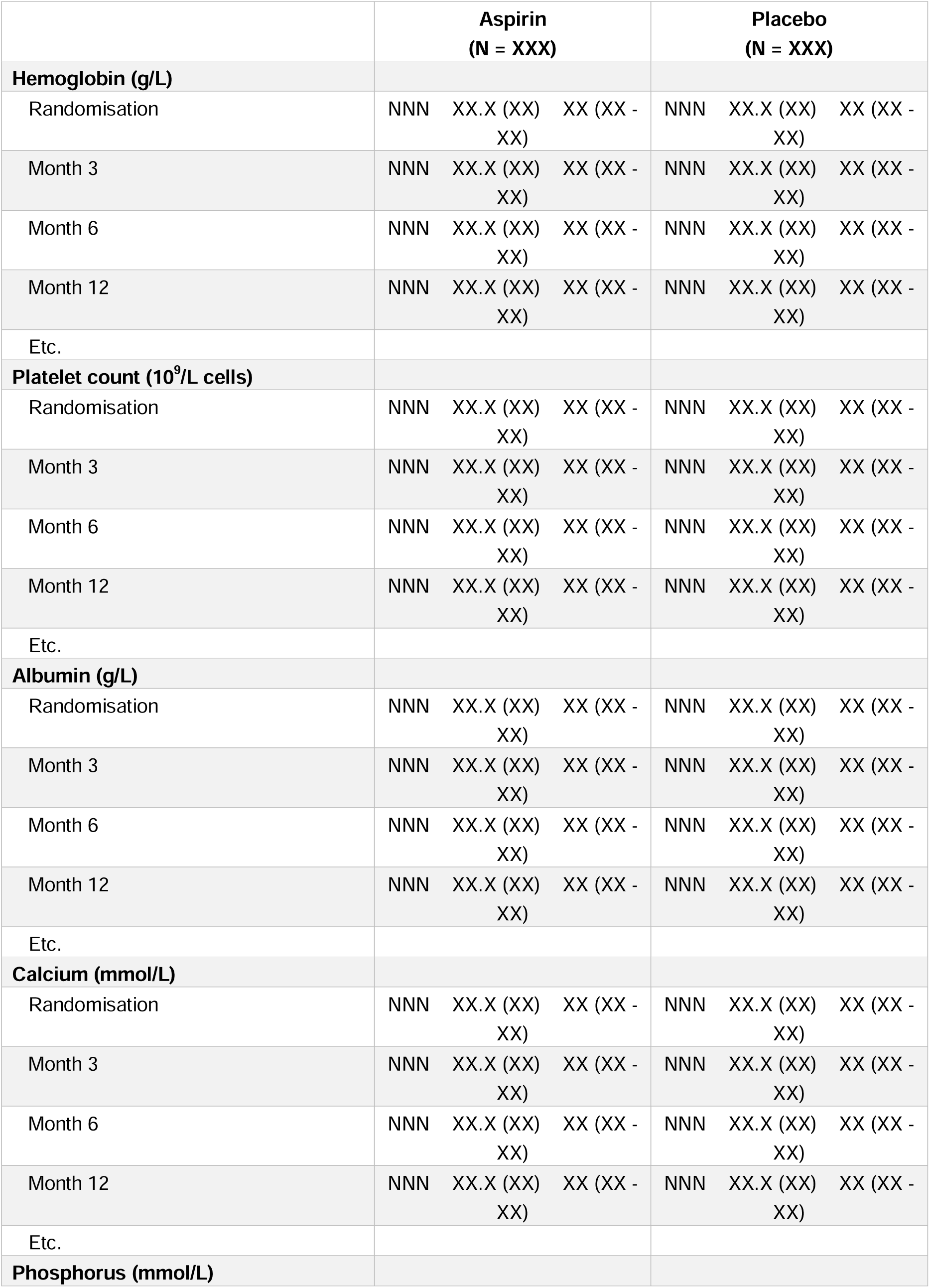
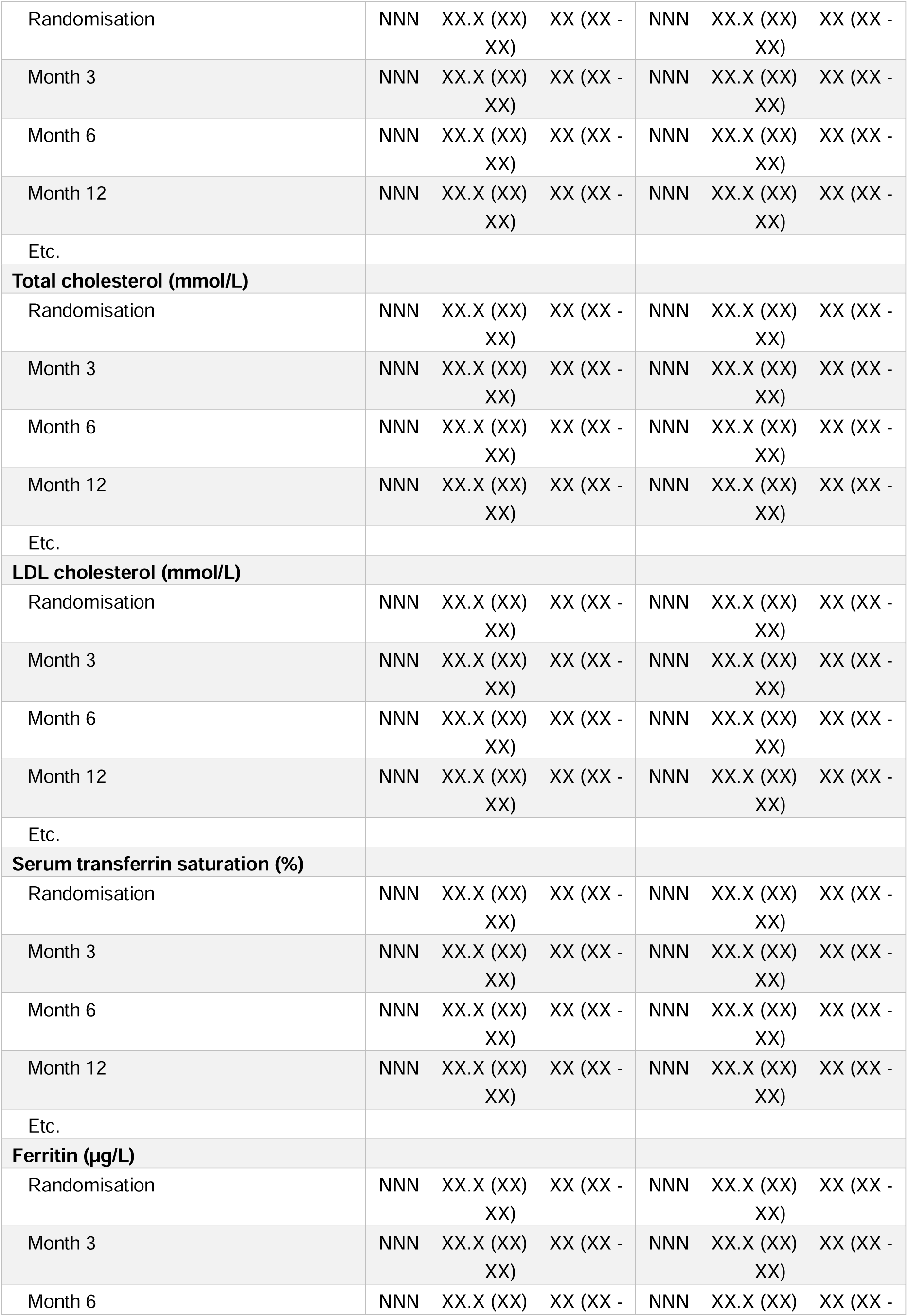
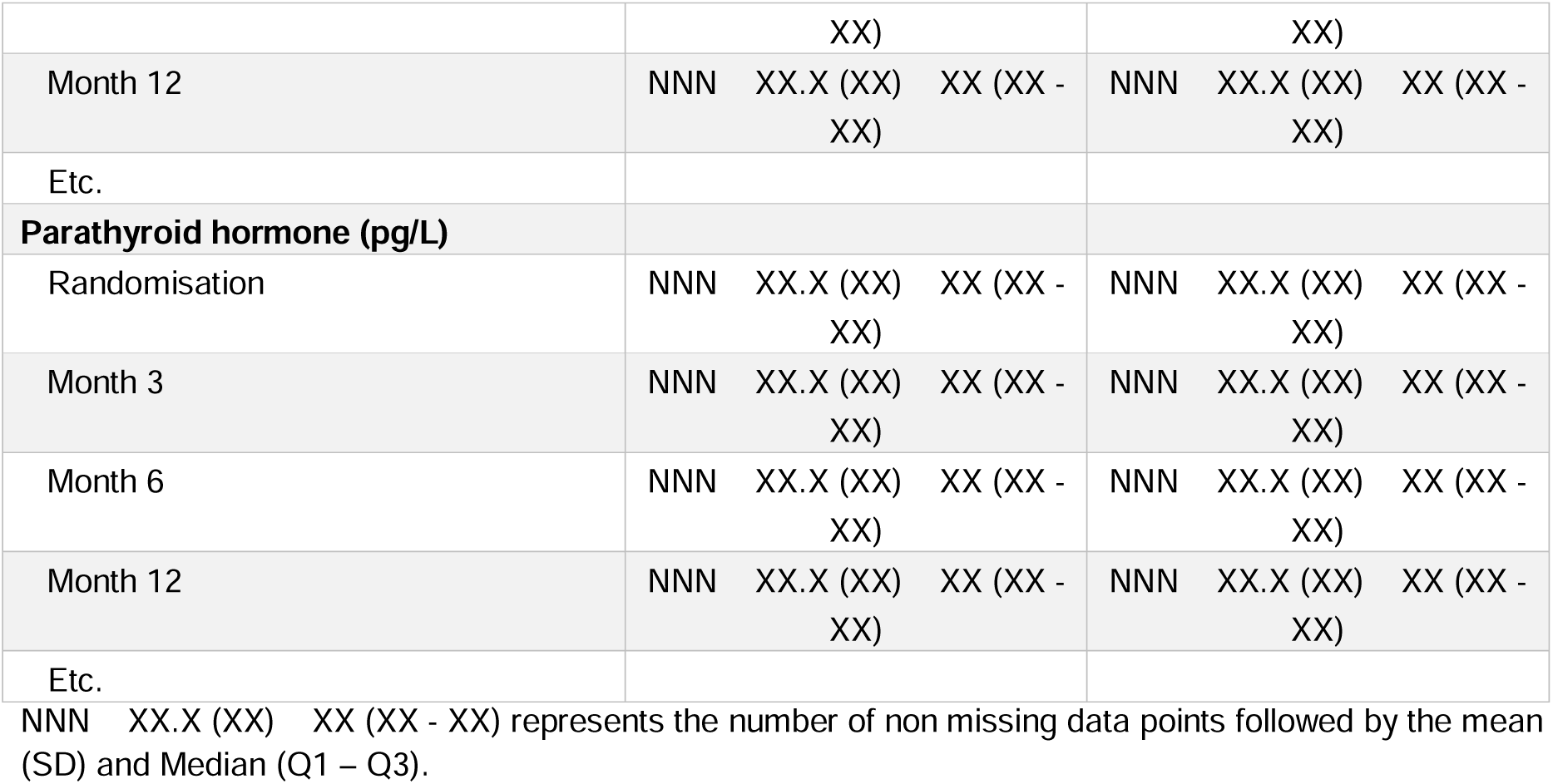
Laboratory results by visit.

|  | Aspirin<br>(N = XXX) |  |  | Placebo<br>(N = XXX) |  |  |
| --- | --- | --- | --- | --- | --- | --- |
| **Hemoglobin (g/L)** |  |  |  |  |  |  |
| Randomisation | NNN | XX.X (XX) | XX (XX - XX) | NNN | XX.X (XX) | XX (XX - XX) |
| Month 3 | NNN | XX.X (XX) | XX (XX - XX) | NNN | XX.X (XX) | XX (XX - XX) |
| Month 6 | NNN | XX.X (XX) | XX (XX - XX) | NNN | XX.X (XX) | XX (XX - XX) |
| Month 12 | NNN | XX.X (XX) | XX (XX - XX) | NNN | XX.X (XX) | XX (XX - XX) |
| Etc. |  |  |  |  |  |  |
| **Platelet count (10<sup>9</sup>/L cells)** |  |  |  |  |  |  |
| Randomisation | NNN | XX.X (XX) | XX (XX - XX) | NNN | XX.X (XX) | XX (XX - XX) |
| Month 3 | NNN | XX.X (XX) | XX (XX - XX) | NNN | XX.X (XX) | XX (XX - XX) |
| Month 6 | NNN | XX.X (XX) | XX (XX - XX) | NNN | XX.X (XX) | XX (XX - XX) |
| Month 12 | NNN | XX.X (XX) | XX (XX - XX) | NNN | XX.X (XX) | XX (XX - XX) |
| Etc. |  |  |  |  |  |  |
| **Albumin (g/L)** |  |  |  |  |  |  |
| Randomisation | NNN | XX.X (XX) | XX (XX - XX) | NNN | XX.X (XX) | XX (XX - XX) |
| Month 3 | NNN | XX.X (XX) | XX (XX - XX) | NNN | XX.X (XX) | XX (XX - XX) |
| Month 6 | NNN | XX.X (XX) | XX (XX - XX) | NNN | XX.X (XX) | XX (XX - XX) |
| Month 12 | NNN | XX.X (XX) | XX (XX - XX) | NNN | XX.X (XX) | XX (XX - XX) |
| Etc. |  |  |  |  |  |  |
| **Calcium (mmol/L)** |  |  |  |  |  |  |
| Randomisation | NNN | XX.X (XX) | XX (XX - XX) | NNN | XX.X (XX) | XX (XX - XX) |
| Month 3 | NNN | XX.X (XX) | XX (XX - XX) | NNN | XX.X (XX) | XX (XX - XX) |
| Month 6 | NNN | XX.X (XX) | XX (XX - XX) | NNN | XX.X (XX) | XX (XX - XX) |
| Month 12 | NNN | XX.X (XX) | XX (XX - XX) | NNN | XX.X (XX) | XX (XX - XX) |
| Etc. |  |  |  |  |  |  |
| **Phosphorus (mmol/L)** |  |  |  |  |  |  |
| Randomisation | NNN | XX.X (XX) | XX (XX - XX) | NNN | XX.X (XX) | XX (XX - XX) |
| Month 3 | NNN | XX.X (XX) | XX (XX - XX) | NNN | XX.X (XX) | XX (XX - XX) |
| Month 6 | NNN | XX.X (XX) | XX (XX - XX) | NNN | XX.X (XX) | XX (XX - XX) |
| Month 12 | NNN | XX.X (XX) | XX (XX - XX) | NNN | XX.X (XX) | XX (XX - XX) |
| Etc. |  |  |  |  |  |  |
| **Total cholesterol (mmol/L)** |  |  |  |  |  |  |
| Randomisation | NNN | XX.X (XX) | XX (XX - XX) | NNN | XX.X (XX) | XX (XX - XX) |
| Month 3 | NNN | XX.X (XX) | XX (XX - XX) | NNN | XX.X (XX) | XX (XX - XX) |
| Month 6 | NNN | XX.X (XX) | XX (XX - XX) | NNN | XX.X (XX) | XX (XX - XX) |
| Month 12 | NNN | XX.X (XX) | XX (XX - XX) | NNN | XX.X (XX) | XX (XX - XX) |
| Etc. |  |  |  |  |  |  |
| **LDL cholesterol (mmol/L)** |  |  |  |  |  |  |
| Randomisation | NNN | XX.X (XX) | XX (XX - XX) | NNN | XX.X (XX) | XX (XX - XX) |
| Month 3 | NNN | XX.X (XX) | XX (XX - XX) | NNN | XX.X (XX) | XX (XX - XX) |
| Month 6 | NNN | XX.X (XX) | XX (XX - XX) | NNN | XX.X (XX) | XX (XX - XX) |
| Month 12 | NNN | XX.X (XX) | XX (XX - XX) | NNN | XX.X (XX) | XX (XX - XX) |
| Etc. |  |  |  |  |  |  |
| **Serum transferrin saturation (%)** |  |  |  |  |  |  |
| Randomisation | NNN | XX.X (XX) | XX (XX - XX) | NNN | XX.X (XX) | XX (XX - XX) |
| Month 3 | NNN | XX.X (XX) | XX (XX - XX) | NNN | XX.X (XX) | XX (XX - XX) |
| Month 6 | NNN | XX.X (XX) | XX (XX - XX) | NNN | XX.X (XX) | XX (XX - XX) |
| Month 12 | NNN | XX.X (XX) | XX (XX - XX) | NNN | XX.X (XX) | XX (XX - XX) |
| Etc. |  |  |  |  |  |  |
| **Ferritin (µg/L)** |  |  |  |  |  |  |
| Randomisation | NNN | XX.X (XX) | XX (XX - XX) | NNN | XX.X (XX) | XX (XX - XX) |
| Month 3 | NNN | XX.X (XX) | XX (XX - XX) | NNN | XX.X (XX) | XX (XX - XX) |
| Month 6 | NNN | XX.X (XX) | XX (XX - XX) | NNN | XX.X (XX) | XX (XX - XX) |

|  |  |  |
| --- | --- | --- |
|  | XX) | XX) |
| Month 12 | NNN XX.X (XX) XX (XX - XX) | NNN XX.X (XX) XX (XX - XX) |
| Etc. |  |  |
| **Parathyroid hormone (pg/L)** |  |  |
| Randomisation | NNN XX.X (XX) XX (XX - XX) | NNN XX.X (XX) XX (XX - XX) |
| Month 3 | NNN XX.X (XX) XX (XX - XX) | NNN XX.X (XX) XX (XX - XX) |
| Month 6 | NNN XX.X (XX) XX (XX - XX) | NNN XX.X (XX) XX (XX - XX) |
| Month 12 | NNN XX.X (XX) XX (XX - XX) | NNN XX.X (XX) XX (XX - XX) |
| Etc. |  |  |

**Table 5.** Patients still on randomised therapy.

| Visit | Aspirin<br>(N = XXX) | Placebo<br>(N = XXX) | Total<br>(N = XXX) |
| --- | --- | --- | --- |
| Month 3 | xx/xxx (xx.x%) | xx/xxx (xx.x%) | xx/xxx (xx.x%) |
| Month 6 | xx/xxx (xx.x%) | xx/xxx (xx.x%) | xx/xxx (xx.x%) |
| Month 12 | xx/xxx (xx.x%) | xx/xxx (xx.x%) | xx/xxx (xx.x%) |
| Etc. for all visits | xx/xxx (xx.x%) | xx/xxx (xx.x%) | xx/xxx (xx.x%) |
| End of study visit * | xx/xxx (xx.x%) | xx/xxx (xx.x%) | xx/xxx (xx.x%) |

\* Used to defined per-protocol analysis set

**Table 6.** Study treatment discontinuations.

| Type of discontinuation / Reason | Aspirin<br>(N = XXX) | Placebo<br>(N = XXX) | Total<br>(N = XXX) |
| --- | --- | --- | --- |
| **Permanent discontinuation** | xx/xxx (xx.x%) | xx/xxx (xx.x%) | xx/xxx (xx.x%) |
| Major bleeding | xx/xxx (xx.x%) | xx/xxx (xx.x%) | xx/xxx (xx.x%) |
| Need for any antiplatelet or anticoagulant drugs due to the condition | xx/xxx (xx.x%) | xx/xxx (xx.x%) | xx/xxx (xx.x%) |
| Other bleeding events that do not meet the diagnostic criteria for major bleeding | xx/xxx (xx.x%) | xx/xxx (xx.x%) | xx/xxx (xx.x%) |
| Primary or secondary end points (excluding non-fatal major bleeding) | xx/xxx (xx.x%) | xx/xxx (xx.x%) | xx/xxx (xx.x%) |
| Patient or investigator determination | xx/xxx (xx.x%) | xx/xxx (xx.x%) | xx/xxx (xx.x%) |
| **Temporary discontinuation** | xx/xxx (xx.x%) | xx/xxx (xx.x%) | xx/xxx (xx.x%) |
| Bleeding episode does not fulfil major bleeding criteria | xx/xxx (xx.x%) | xx/xxx (xx.x%) | xx/xxx (xx.x%) |
| Gastrointestinal symptoms | xx/xxx (xx.x%) | xx/xxx (xx.x%) | xx/xxx (xx.x%) |
| Surgery or invasive procedure | xx/xxx (xx.x%) | xx/xxx (xx.x%) | xx/xxx (xx.x%) |
| Participant's or physician's decision | xx/xxx (xx.x%) | xx/xxx (xx.x%) | xx/xxx (xx.x%) |
| Other | xx/xxx (xx.x%) | xx/xxx (xx.x%) | xx/xxx (xx.x%) |

**Table 7.**
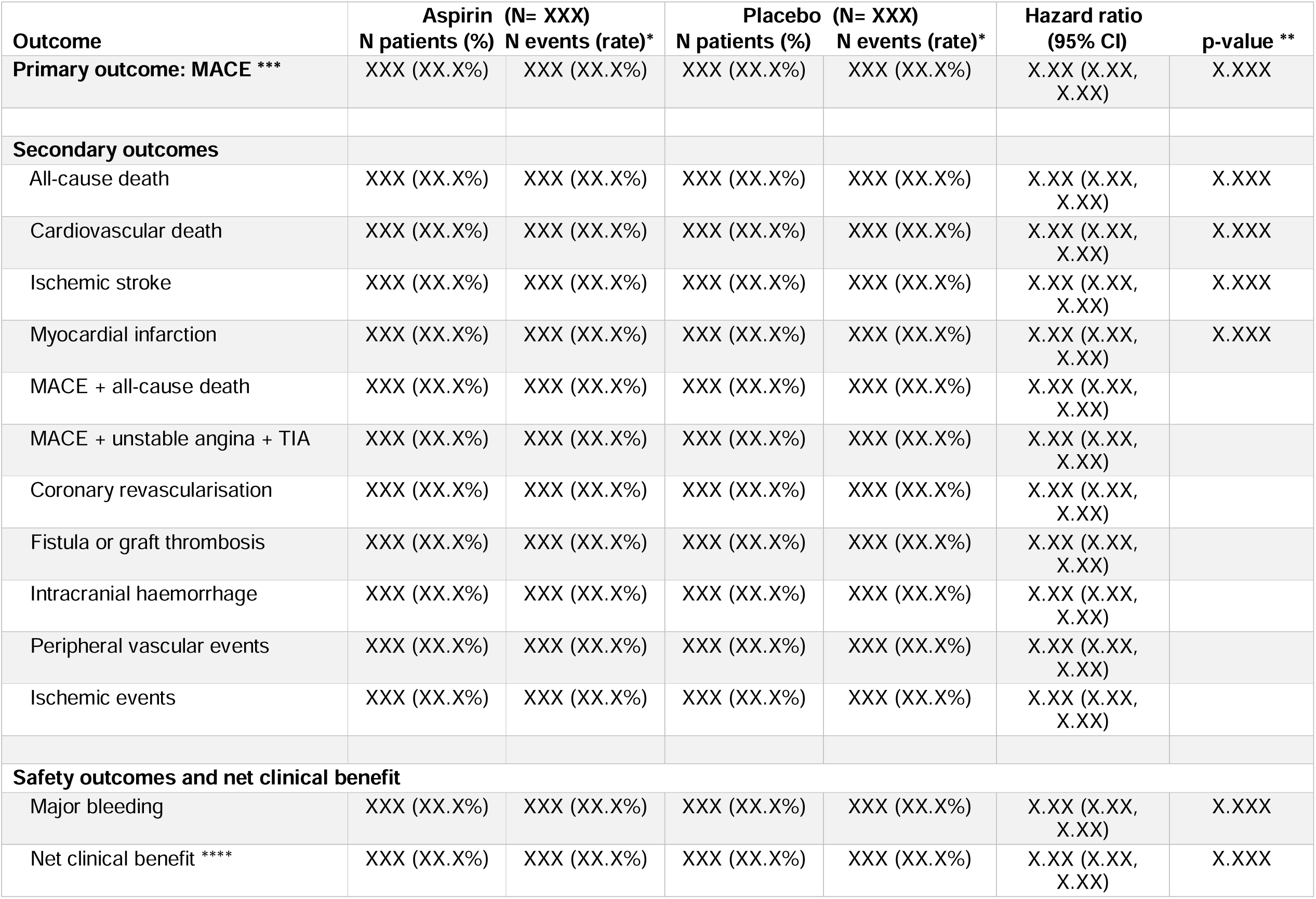
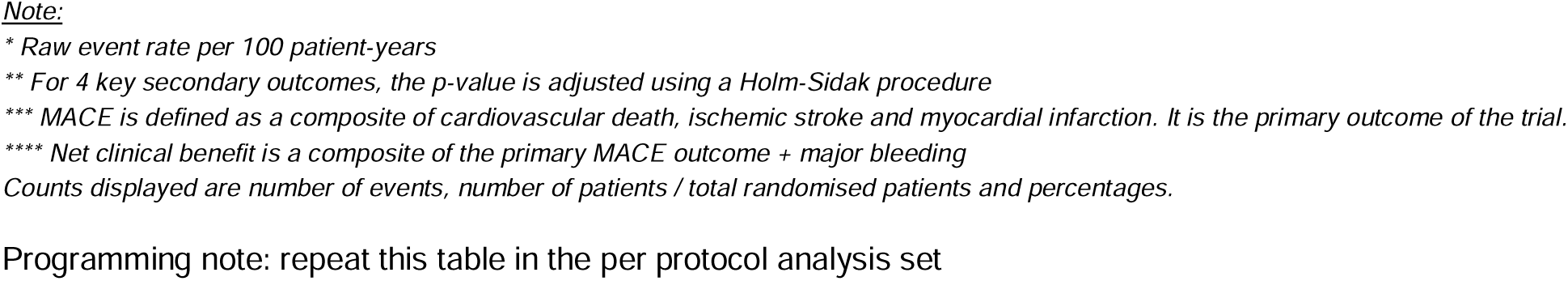
Survival analysis of primary and secondary outcomes.

| **Outcome** | **Aspirin (N= XXX)** |  | **Placebo (N= XXX)** |  | **Hazard ratio (95% CI)** | **p-value \*\*** |
| --- | --- | --- | --- | --- | --- | --- |
|  | **N patients (%)** | **N events (rate)\*** | **N patients (%)** | **N events (rate)\*** |  |  |
| **Primary outcome: MACE \*\*\*** | XXX (XX.X%) | XXX (XX.X%) | XXX (XX.X%) | XXX (XX.X%) | X.XX (X.XX, X.XX) | X.XXX |
| **Secondary outcomes** |  |  |  |  |  |  |
| All-cause death | XXX (XX.X%) | XXX (XX.X%) | XXX (XX.X%) | XXX (XX.X%) | X.XX (X.XX, X.XX) | X.XXX |
| Cardiovascular death | XXX (XX.X%) | XXX (XX.X%) | XXX (XX.X%) | XXX (XX.X%) | X.XX (X.XX, X.XX) | X.XXX |
| Ischemic stroke | XXX (XX.X%) | XXX (XX.X%) | XXX (XX.X%) | XXX (XX.X%) | X.XX (X.XX, X.XX) | X.XXX |
| Myocardial infarction | XXX (XX.X%) | XXX (XX.X%) | XXX (XX.X%) | XXX (XX.X%) | X.XX (X.XX, X.XX) | X.XXX |
| MACE + all-cause death | XXX (XX.X%) | XXX (XX.X%) | XXX (XX.X%) | XXX (XX.X%) | X.XX (X.XX, X.XX) |  |
| MACE + unstable angina + TIA | XXX (XX.X%) | XXX (XX.X%) | XXX (XX.X%) | XXX (XX.X%) | X.XX (X.XX, X.XX) |  |
| Coronary revascularisation | XXX (XX.X%) | XXX (XX.X%) | XXX (XX.X%) | XXX (XX.X%) | X.XX (X.XX, X.XX) |  |
| Fistula or graft thrombosis | XXX (XX.X%) | XXX (XX.X%) | XXX (XX.X%) | XXX (XX.X%) | X.XX (X.XX, X.XX) |  |
| Intracranial haemorrhage | XXX (XX.X%) | XXX (XX.X%) | XXX (XX.X%) | XXX (XX.X%) | X.XX (X.XX, X.XX) |  |
| Peripheral vascular events | XXX (XX.X%) | XXX (XX.X%) | XXX (XX.X%) | XXX (XX.X%) | X.XX (X.XX, X.XX) |  |
| Ischemic events | XXX (XX.X%) | XXX (XX.X%) | XXX (XX.X%) | XXX (XX.X%) | X.XX (X.XX, X.XX) |  |
| **Safety outcomes and net clinical benefit** |  |  |  |  |  |  |
| Major bleeding | XXX (XX.X%) | XXX (XX.X%) | XXX (XX.X%) | XXX (XX.X%) | X.XX (X.XX, X.XX) | X.XXX |
| Net clinical benefit **** | XXX (XX.X%) | XXX (XX.X%) | XXX (XX.X%) | XXX (XX.X%) | X.XX (X.XX, X.XX) | X.XXX |

Note:
\* Raw event rate per 100 patient-years
\*\* For 4 key secondary outcomes, the p-value is adjusted using a Holm-Sidak procedure
\*\*\* MACE is defined as a composite of cardiovascular death, ischemic stroke and myocardial infarction. It is the primary outcome of the trial.
\*\*\*\* Net clinical benefit is a composite of the primary MACE outcome + major bleeding
Counts displayed are number of events, number of patients / total randomised patients and percentages.
Programming note: repeat this table in the per protocol analysis set

**Table 8.** Major bleeding events.

| Outcome | Aspirin<br>(N= XXX)<br>Ev Pt (%) | Placebo<br>(N= XXX)<br>Ev Pt (%) | Fisher's<br>exact test<br>P-value |
| --- | --- | --- | --- |
| **Major bleeding** | XXX XXX (XX.X%) | XXX XXX (XX.X%) | **0.xxx** |
| Fatal bleeding | XXX XXX (XX.X%) | XXX XXX (XX.X%) |  |
| Symptomatic bleeding into a critical area<br>or bleeding into the surgical site requiring<br>reoperation | XXX XXX (XX.X%) | XXX XXX (XX.X%) |  |
| Bleeding requiring hospitalisation | XXX XXX (XX.X%) | XXX XXX (XX.X%) |  |
| Bleeding requiring transfusion |  |  |  |
| **Fatal bleeding + intracranial bleeding** | XXX XXX (XX.X%) | XXX XXX (XX.X%) | **0.xxx** |
| **Site of bleeding** |  |  |  |
| Gastrointestinal | XXX XXX (XX.X%) | XXX XXX (XX.X%) |  |
| Intracranial | XXX XXX (XX.X%) | XXX XXX (XX.X%) |  |
| Intraocular | XXX XXX (XX.X%) | XXX XXX (XX.X%) |  |
| Respiratory | XXX XXX (XX.X%) | XXX XXX (XX.X%) |  |
| Abdominal | XXX XXX (XX.X%) | XXX XXX (XX.X%) |  |
| Retroperitoneal | XXX XXX (XX.X%) | XXX XXX (XX.X%) |  |
| Other site | XXX XXX (XX.X%) | XXX XXX (XX.X%) |  |

Notes:
Counts displayed are number of events, number of patients / total randomised patients and percentages.
Site of bleeding includes major and non-major bleeding.
Programming note: repeat this table in per protocol analysis set

**Table 9.**
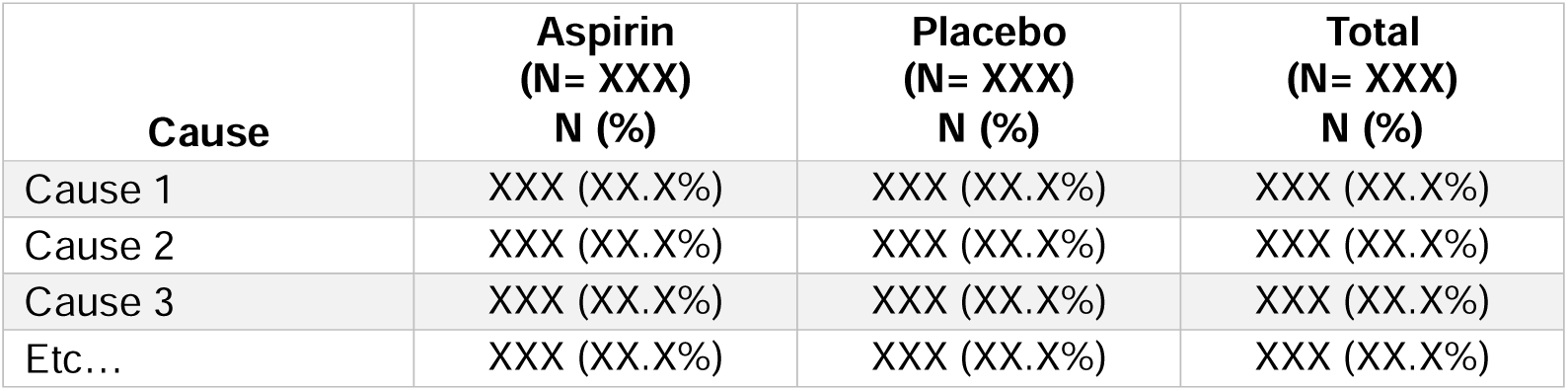
Causes of deaths.

## Data Availability

Not relevant

